# A Residual Approach to Estimate Biological Age from Gompertz Modeling

**DOI:** 10.1101/2025.11.06.25339716

**Authors:** Hui Zhang, Shuishan Zhang, Xilu Wang, Yaqi Huang, Kuaile Dong, Xianglin Aneta Guo, Xueqin Li, Shuai Jiang, Zixin Hu, Xiaofeng Wang, Jie Chen, Zhijun Bao, Jiucun Wang, Li Jin, Yiqin Huang, Yi Li, Meng Hao

## Abstract

Biological age (BA) and its residual relative to chronological age are popularly used to quantify individual aging. Although these residuals independently predict age-related health outcomes, conventional BA measures often lack robustness in heterogeneous populations, and their residuals are not directly derivable in clinical practice. To address these limitations, we introduce the Gompertz law-based residual (GOLD-R) framework, a method designed to directly estimate BA residuals and optimized for cross-sectional data. We demonstrated the applicability and robustness of GOLD-R across multiple data types and populations. First, training on DNA methylation data from the EWAS Data Hub, the framework outperformed established epigenetic clocks in predicting mortality in a pan-cancer dataset. Then, applied to UK Biobank proteomics data, GOLD-R generated organismal and organ-specific aging measures that proved more robust than conventional age-prediction approaches in forecasting incident diseases and mortality. Finally, extending the analysis to clinical biomarkers using the NHANES and HRS, we found that GOLD-R residuals, derived from clinical biomarkers, surpassed those from both epigenetic and phenotypic clocks in performance. In summary, our findings establish GOLD-R as a robust algorithm for biological age estimation, providing a practical tool for both research and clinical applications.

## Introduction

Biological age (BA), a metric for quantifying the aging process, has garnered increasing interest for its potential to reflect aging status more accurately than chronological age (CA) ^1^. It refers to age as determined by biological metrics, typically constructed from clinical or omics (e.g, epigenetics, proteomics) indicators^2^. It more accurately reflects an individual’s actual state of aging. Early first-generation models (e.g., HorvathAge, KDM Age) directly predicted chronological age (CA)^3,4^. In contrast, second-generation models (e.g., GrimAge, LevineAge) are considered more practical as they predict mortality risk and other aging factors^5,6^. While second-generation BA models often depend on longitudinal data for training, they might lack robustness and reproducibility in external validation due to heterogeneity among populations^7^. This progression raises a key question: can the construction of BA be further optimized?

Beyond BA itself, its residual in relation to CA is a clinically informative measure of an individual’s aging^8,9^.It was typically minimal in healthy populations but became pronounced in individuals with diseases or accelerated aging^10,11^. This measure independently predicts health outcomes, capturing risk beyond CA alone. For instance, Oh et al. utilized age gap measurements to quantify aging status relative to same-aged peers and established associations with organ-specific age-related diseases^12,13^. Furthermore, current aging clocks calculate their residuals, or "acceleration" through linear regression, which limits their direct derivation in clinical settings^14^. Thus, we hypothesized that enabling biomarkers to directly predict this deviation would refine BA estimation and enhance clinical utility. This strategy could provide an intuitive interpretation by directly linking biomarker profiles to an individual’s actual state of aging relative to their peers. Previously, we proposed a BA model based on the Gompertz law based on CA and biomarkers, termed GOLD BioAge, and revealed its effective prediction for mortality and disease risk^11^. The model employed a residual-based approach, using biomarkers to quantify the mortality risk not captured by CA. This risk was converted into a composite measure of deviation from CA, expressed in years, which was then added to CA to derive an individual’s BA. In essence, the model estimated the residual of BA relative to CA to assess individual’s actual state of aging. Although this approach demonstrated effectiveness in longitudinal cohorts, the limited availability of high-quality longitudinal data represents a significant constraint for widespread application.

Here, we propose a residual-based framework optimized for cross-sectional data, which represents the most common scenario in clinical practice and research settings. This Gompertz law-based (GOLD-R) approach aims to derive aging clocks by directly predicting biological age residuals. First, to establish the method’s validity in epigenetic aging, we developed GOLD-R DNAmAge using DNA methylation data from the EWAS Data Hub. This epigenetic clock was validated in external datasets. Next, we assessed the framework’s performance using proteomics data from the UK Biobank to predict age-related health outcomes. Finally, to demonstrate translational potential in clinically accessible measures, we applied GOLD-R to standard clinical biomarkers from the U.S. NHANES and HRS. The resulting aging clocks were benchmarked against established epigenetic and phenotypic measures, confirming the framework’s robustness across data types.

## Results

### The concept of GOLD-R framework

This Gompertz law-based residual approach (GOLD-R) posits that mortality risk increases exponentially with chronological age (CA). Individual deviations from this general trend are termed residual risks. (**Figure 1A-B**). It involves a two-stage modeling process to derive the residuals (**Figure 1C**). First, we generate an initial residual representing the discrepancy between the CA-based Gompertzian risk and the risk predicted by the biomarkers. This initial residual is then refined through a second, distinct model where the rest biomarkers are used to predict the residual itself, capturing a biologically informed signal of aging status. Finally, this refined residual is converted into year-equivalent units and added to the CA to yield the final GOLD-R BioAge, providing an intuitive measure of personalized aging. The comprehensive workflow is illustrated in **Figure 1 and S1**.

**Figure 1.**
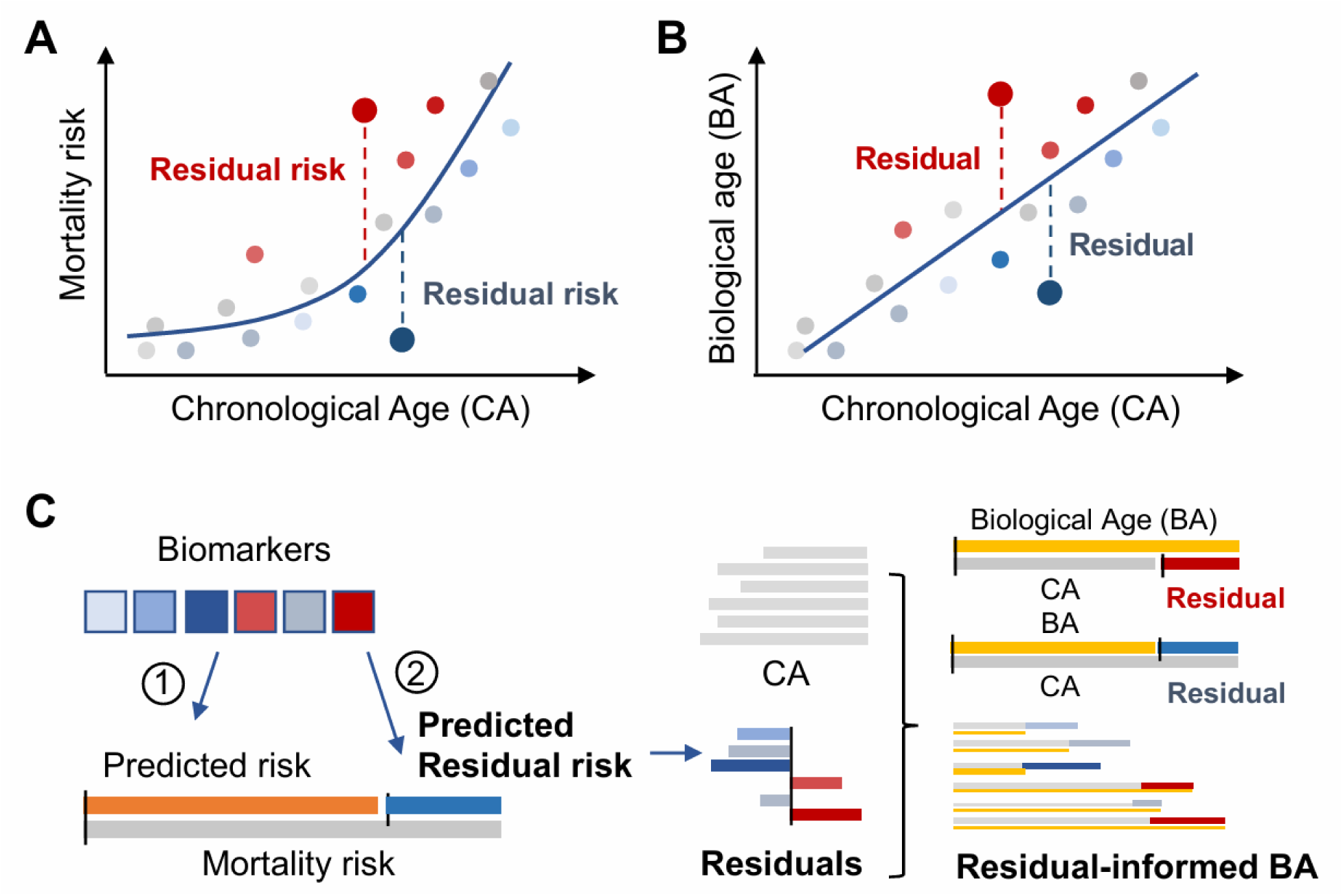
Schematic diagram of the GOLD-R BioAge computational framework. The diagram (A) illustrated the exponential relationship between mortality risk and chronological age (CA). “Residual risk” denotes the discrepancy between mortality risk attributable to biological characteristics and the risk expected for the individual’s chronological age, where red indicates accelerated ageing and blue denotes decelerated ageing. The panel (B) showed the relationship between biological age (BA), chronological age (CA), and residual. Diagram (C) depicted the core algorithm of GOLD-R Age: first, biomarkers were utilized to predict mortality risk, yielding the residual risk. This residual was then predicted by the biomarkers. Finally, the residual was combined with chronological age to derive the residual-informed biological age.

### Development and validation of GOLD-R DNAmAge and benchmark analysis

We first applied this algorithm on DNA methylation (DNAm) data from EWAS Data Hub. During the training phase, DNAm data (across 21 tissues and cell types) from 7,313 samples (mean age: 56.05 ± 19.46 years) were profiled and analyzed using Illumina 450K arrays. We developed GOLD-R DNAmAge through a two-step LASSO regression approach: the first model (615 CpGs) estimated mortality risk and generated residual risk scores, while the second model (34 CpGs) predicted these residuals, collectively constituting the GOLD-R DNAmAge algorithm.

For validation, we assessed its prognostic utility in DNAm data of pan-cancer tissues, comprising 5,502 participants (mean age: 59.14 ± 15.05 years). Over a median follow-up of 1.92 years (IQR: 1.05-3.75), 1,815 deaths were documented. As shown in **Figure 2A**, GOLD-R DNAmAge and its residual component demonstrated robust age correlations across both training and validation datasets. In comparative analyses, we evaluated multiple established epigenetic clocks (**Table 1**). Correlation analysis (**Figure 2B**) revealed strong associations among first-generation clocks (such as BNN, HorvathAge, HannumAge, DNAmAge) and among second-generation clocks (such as GrimAge, GOLD-R DNAmAge), while DunedinPACE emerged as a distinct indicator of aging pace.

**Figure 2.**
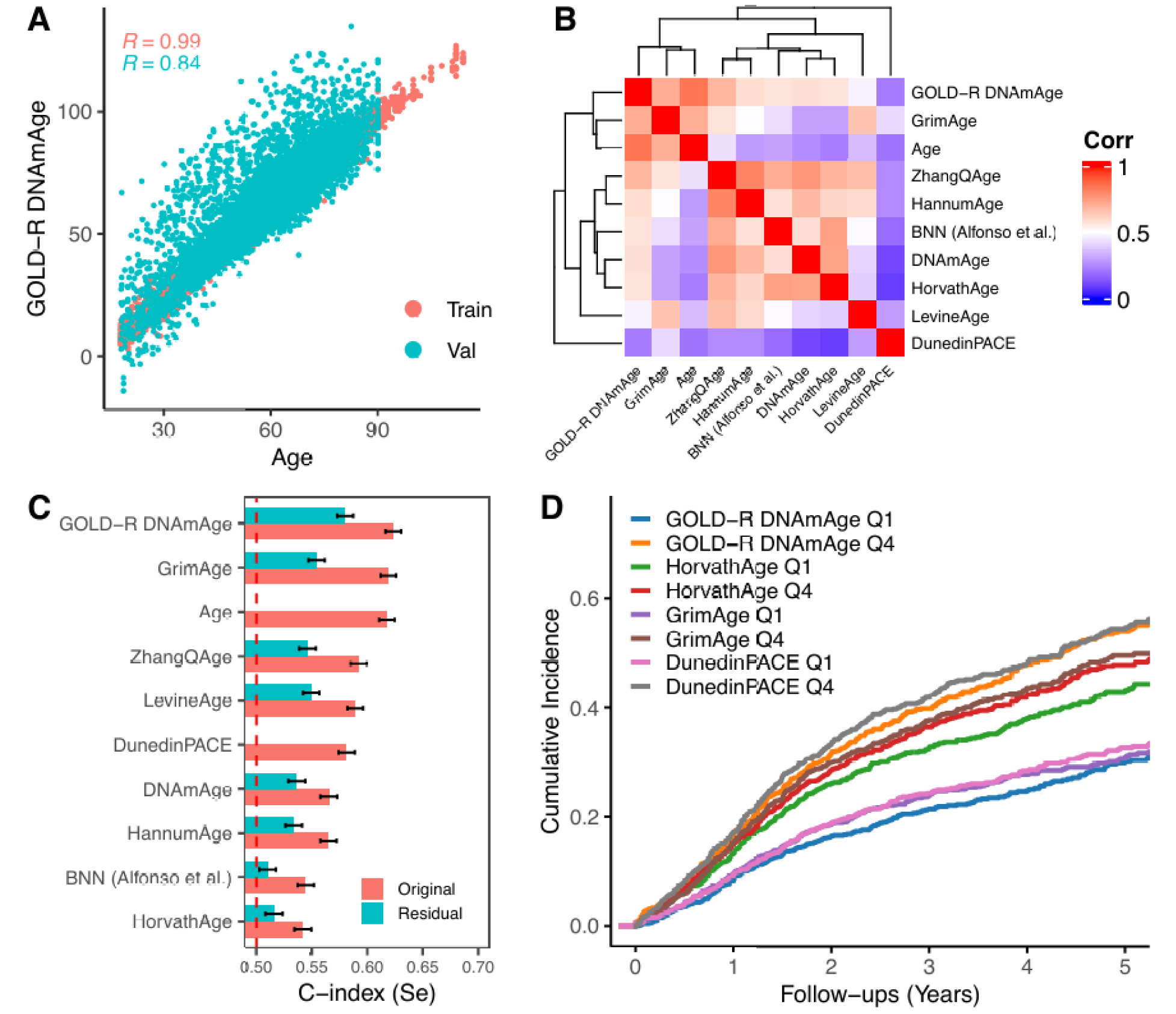
The associations of GOLD DNAmAge with mortality. Scatter plots (A) demonstrated the correlation between GOLD DNAmAge and chronological age in train and validation (Val) datasets. The heatmap (B) filled with Pearson correlations among epigenetic clocks. The bar chart (C) compared C-index values for predicting mortality risk across clocks (and their residuals). Survival curves (D) of lowest and highest quartile group (Q1, Q4) based on GOLD-R DNAmAge and other clocks in 5-year mortality of cancer prognosis.

**Table 1.**
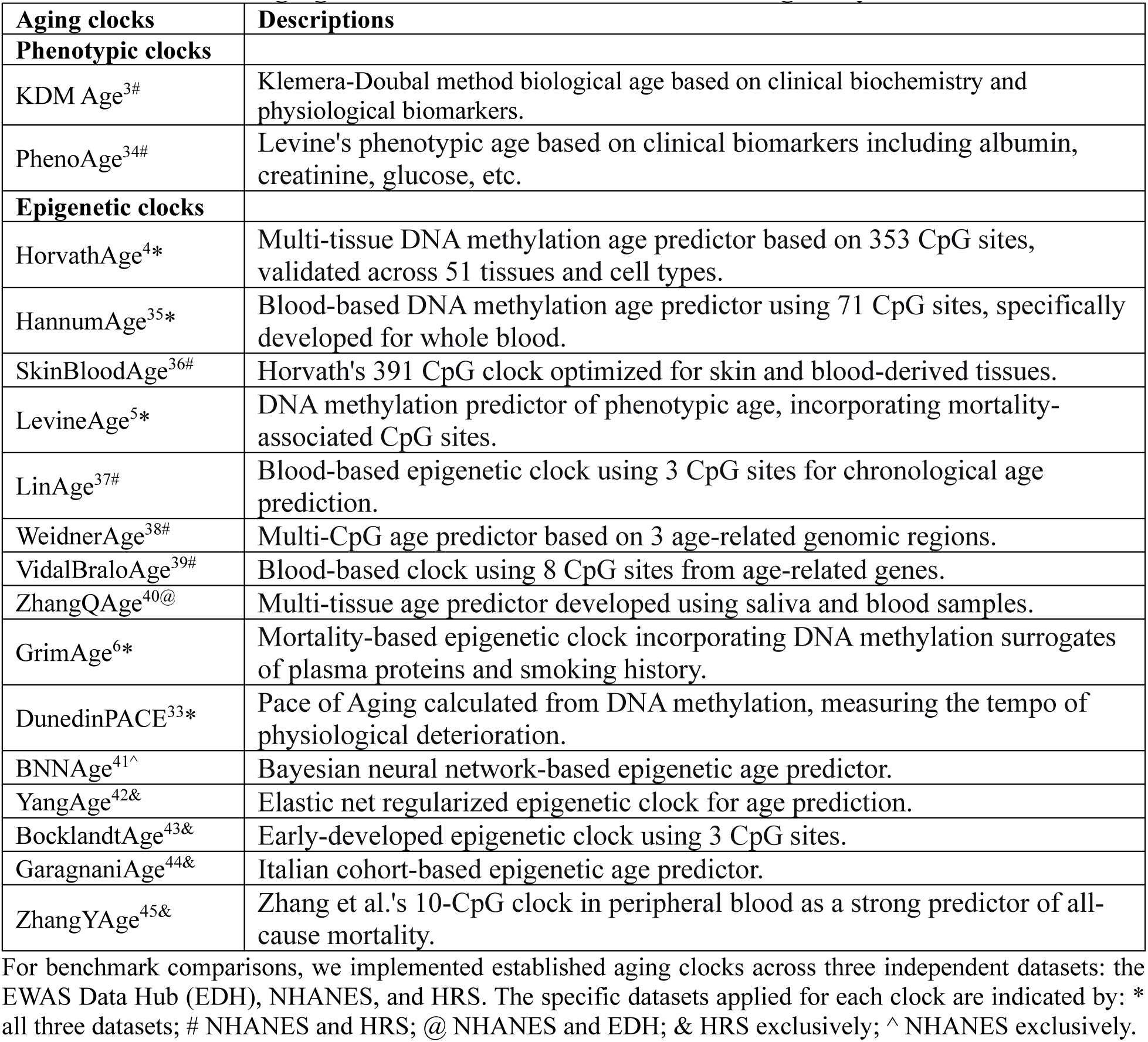
Established aging clocks included in the benchmarking analysis.

GOLD-R DNAmAge achieved one of the highest C-index values for predicting mortality risk (**Figure 2C**). Following adjustment for age, sex, and race, GOLD-R DNAmAge demonstrated a significant association with all-cause mortality (adjusted hazard ratio [aHR]: 1.013, 95% confidence interval [CI]: 1.008-1.017, p<0.01, **Figure S3A**). Several established epigenetic clocks also showed significant mortality associations: HannumAge (aHR: 1.003, 95% CI: 1.001-1.005), LevineAge (aHR: 1.003, 95% CI: 1.002-1.004), BNN (aHR: 1.003, 95% CI: 1.001-1.006), ZhangQAge (aHR: 1.005, 95% CI: 1.003-1.007), GrimAge (aHR: 1.015, 95% CI: 1.011-1.019), and DunedinPACE (aHR: 1.075, 95% CI: 1.057-1.093). In contrast, neither DNAmAge (aHR: 1.001, 95% CI: 0.999-1.003) nor HorvathAge (aHR: 1.001, 95% CI: 0.999-1.003) reached statistical significance.

GOLD-R DNAmAge and its residual exhibited superior discriminative performance for mortality prediction (C-index: 0.623, 0.580) compared to other epigenetic clocks, including GrimAge (0.619, 0.554), LevineAge (0.589, 0.549), DunedinPACE (0.581), and first-generation clocks (**Figure 2C, S3B**). In analyses of 5-year cancer mortality (**Figure 2D**), the highest quartile of GOLD-R DNAmAge residuals was associated with a mortality rate of 69.9%, substantially higher than the lowest quartile (46.0%). The corresponding mortality rate of highest-risk group were 56.9% for HorvathAge, 69.1% for GrimAge, and 67.4% for DunedinPACE. Collectively, these findings underscore the enhanced capability of GOLD-R DNAmAge to capture mortality risk.

### Applications of GOLD-R on plasma proteomics

To extend the applicability, we applied our framework to plasma proteomics from the UK Biobank (UKB), deriving a novel measure termed GOLD-R ProtAge, at both the organismal and organ-specific levels (**Figure S4**). The analysis encompassed 53,014 participants (mean age 56.81 ± 8.21 years) and 2,923 protein biomarkers. For comparison, a conventional proteomic-based biological age (ProtAge) was also constructed using a direct age-prediction approach. All models were developed and evaluated under a 5-fold cross-validation scheme, with performance metrics assessed on held-out test sets.

GOLD-R ProtAge demonstrated significantly improved predictive accuracy for all-cause mortality compared to conventional ProtAge (**Figure 3A**). This was reflected in a higher adjusted hazard ratio (aHR = 1.177, 95% CI: 1.168–1.185 vs. aHR = 1.085, 95% CI: 1.079–1.092) and a substantially better concordance index (C-index = 0.740 vs. 0.588). The superiority of GOLD-R ProtAge was consistently observed across various cause-specific mortality endpoints. In predicting incident chronic diseases, GOLD-R ProtAge also outperformed the conventional model across all conditions examined (**Figure 3A**, right panel). Notably, it exhibited strong discriminatory performance for dementia (C-index = 0.773) and COPD (C-index = 0.719), markedly surpassing the corresponding ProtAge estimates (C-indices = 0.574 and 0.567, respectively).

**Figure 3.**
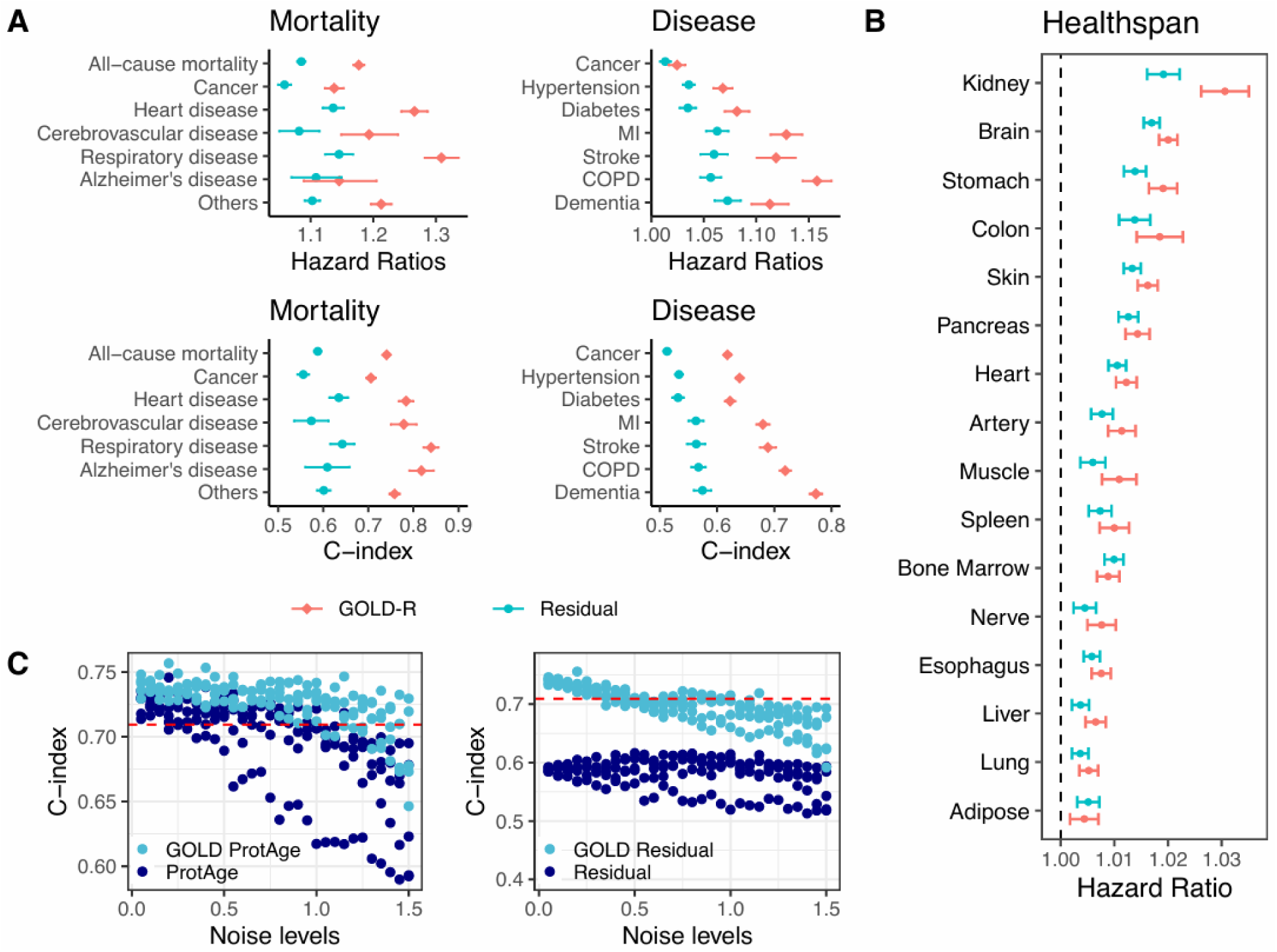
Predictive performance of GOLD-R residuals for mortality and disease risk in the UKB. (A) Forest plots displayed adjusted hazard ratios (adjusting covariates) and C-index values (crude models) for GOLD-R residuals in relation to all-cause mortality, cause-specific mortality, and incident diseases. (B) Forest plots of adjusted hazard ratios (aHR) of organ-specific GOLD-R ProtAge for predicting healthspan. The aHR of GOLD-R was marked in red, and conventional ProtAge residual was marked in blue. (C) Trend analysis shows how C-index values change with the incremental addition of normally distributed noise to test datasets.

The resulting organ-specific GOLD-R ProtAge models showed enhanced performance in predicting healthspan compared to their conventional ProtAge counterparts (**Figure 3B**). Among these, the kidney (aHR = 1.031, 95% CI: 1.022–1.040), brain (aHR = 1.020, 95% CI: 1.013–1.027), and stomach (aHR = 1.019, 95% CI: 1.011–1.027) models exhibited the strongest associations with healthspan outcomes (**Figure S5**). To evaluate model robustness, we conducted sensitivity analyses by introducing normally distributed noise into the test datasets, simulating potential population differences between training and validation contexts (**Figure 3C**). GOLD-R ProtAge maintained exceptional discrimination performance across all noise levels, consistently outperforming the conventional ProtAge model under these simulated heterogeneous conditions.

### Predicting Epigenetic Residuals with Clinical Biomarkers

We validated the residual-informed GOLD-R framework using data from NHANES 1999-2002 (N=2,532; mean age 66.1±10.1 years; 1,361 deaths over 17.2 years) and HRS 2016 (N=4,018; mean age 69.9±9.5 years; 766 deaths over 7.1 years). Due to unavailability of raw DNAm data, we derived aging residuals from established epigenetic clocks and modeled them against clinical biomarkers to construct GOLD-R DNAmCliAge. There were strong correlations between GOLD- R DNAmCliAge and CA in the NHANES and HRS, indicating robust generalization capabilities (**Figure 4A-B**). In NHANES, GOLD-R DNAmCliAge showed competitive mortality prediction (C-index=0.75), comparable to GrimAge (0.76) and superior to first-generation clocks. Its age-adjusted residual component demonstrated stronger discrimination (C-index=0.64) versus GrimAge (0.61), PhenoAge (0.56), and DunedinPACE (0.59). Similarly, in HRS, GOLD-R DNAmCliAge achieved the highest performance (C-index=0.77) among all clocks, with its residual component (C-index=0.62) outperforming alternatives (**Figure 4C-D**). The superiority of GOLD-R in capturing mortality risk was confirmed by the distinct separation of survival curves for its high-risk and low-risk groups, defined by model residuals (**Figure 5A-B**). For example, the top quartile of the GOLD-R residual was associated with a 10-year mortality rate of 55.3%, which is lower than the rates for GrimAge (60.0%) and HorvathAge (65.1%).

**Figure 4.**
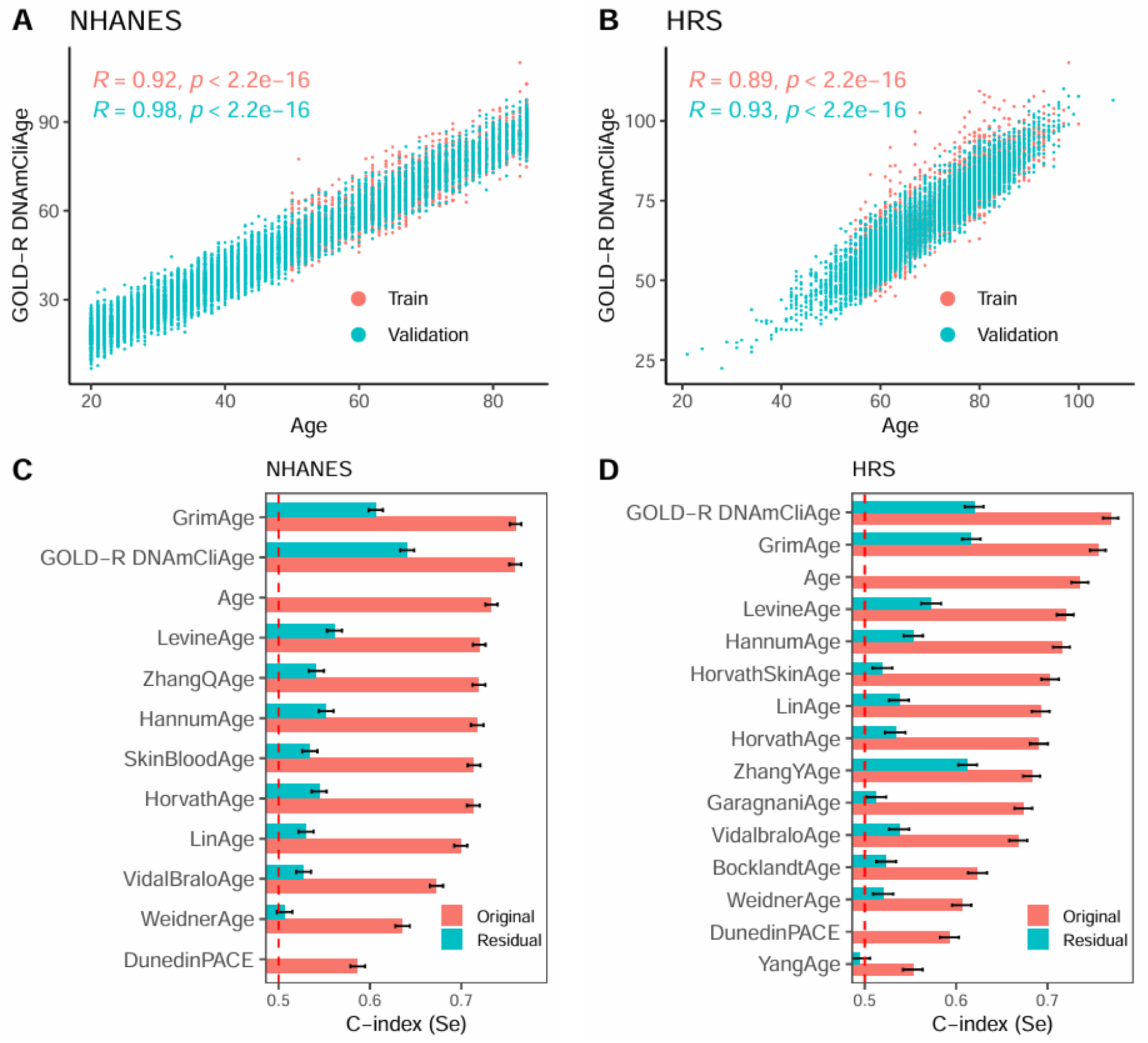
GOLD-R DNAmCliAge model in the NHANES and HRS cohorts. Scatter plots (A-B) demonstrated the strong correlations between GOLD-R DNAmCliAge and chronological age in the NHANES and HRS cohorts, respectively. Pearson correlation coefficients were presented. (C-D) illustrated the C-index comparison of GOLD-R DNAmCliAge and its residual with other epigenetic clocks in mortality prediction.

**Figure 5.**
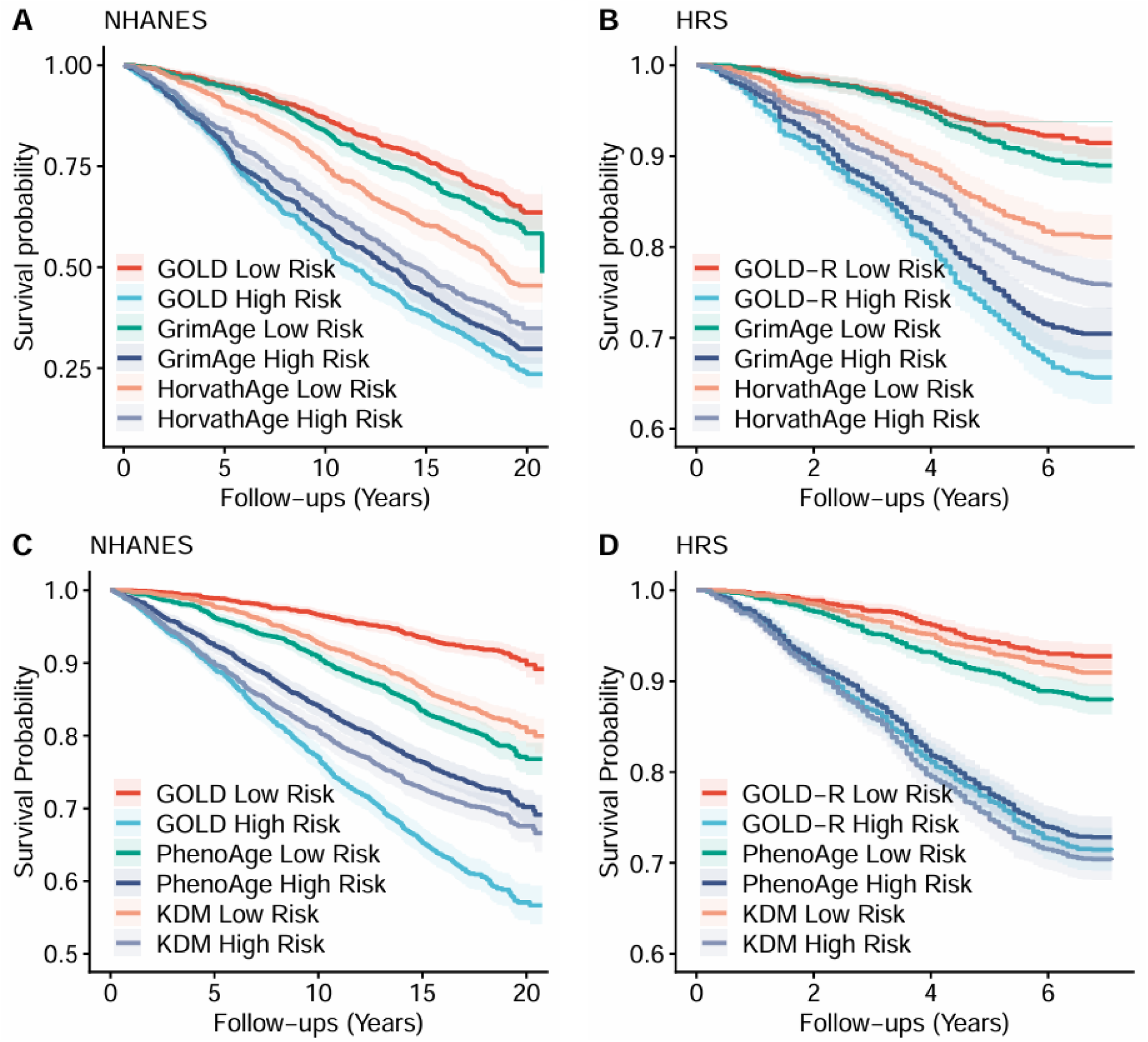
Associations of GOLD DNAmCliAge with mortality risk in NHANES and HRS. Survival curves stratified by residuals of GOLD-R, GrimAge and HorvathAge (A, B) from NHANES and HRS cohorts (training groups), and individual survival curves stratified by GOLD-R, PhenoAge and KDM (C, D) from both cohorts (validation groups). The high-risk and low-risk groups represented the top and bottom 25% of individual residual, respectively, within each biological age stratification. KDM: Klemera-Doubal method.

Independent validation in NHANES (N=8,803; 1,825 deaths) and HRS (N=5,958; 968 deaths) confirmed these findings (**Figure 5C-D**). GOLD-R residuals maintained superior prediction (NHANES: 0.70; HRS: 0.67) versus PhenoAge (0.59, 0.66) and KDM Age (0.54, 0.61). For instance, in NHANES, the highest-risk quartile of GOLD-R residual showing the lowest 10-year survival probability (77.0% vs 80.7% for PhenoAge and 84.1% for KDM Age) **(Figure 5C**). Collectively, GOLD-R DNAmCliAge demonstrated consistent superiority across two cohorts, establishing it as a robust mortality risk predictor.

## Discussions

This study introduced a novel residual-based framework that integrated the Gompertz law of mortality to quantify aging status. We validated this approach across three complementary data modalities in four independent cohorts, proceeding from established epigenetic measures to proteomic applications and finally to clinical translation. Our benchmark analysis included fifteen established epigenetic clocks (**Table 1**), and two phenotypic clocks for comparison. First, Using DNA methylation data, we developed the cross-tissue GOLD-R DNAmAge, and it demonstrated superior mortality prediction compared to first-generation epigenetic clocks and achieved performance comparable to GrimAge. When applied to plasma proteomics from UK Biobank, our residual-based approach surpassed conventional age prediction in forecasting incident age-related diseases and mortality. Additionally, it enabled BA estimation for multiple organ systems through organ-specific protein signatures. Extending our validation to clinical biomarkers, we developed a GOLD-R DNAmCliAge measure by predicting DNAm-derived residual risks. This approach yielded a metric that surpassed both Levine’s PhenoAge and KDM Age in predictive accuracy. In summary, these results validate the efficacy of the GOLD-R framework in constructing accurate aging clocks through residual prediction, establishing its utility across diverse biological data modalities.

Previous studies reported that GrimAge outperformed other aging clocks in predicting mortality among general population, including CA, HorvathAge, HannumAge, and PhenoAge^15–17^. For instance, GrimAge was reported as the strongest predictor of mortality among all DNAm clocks and CA in NHANES^15^. In addition, the potential ability of various BAs for prognostic prediction was also investigated in cancer survivors^18,19^. Guida et.al included 4117 cancer survivors (mean age: 35.08 [1.17] years) and calculated seven BAs in the St. Jude Lifetime (SJLIFE) Cohort. They reported that BA acceleration was consistently and robustly associated with all-cause mortality, such as HorvathAge, HannumAge, PhenoAge, and GrimAge acceleration^18^. Notably, survivors with the highest GrimAge acceleration (HR 7.59, 95% CI: 3.36-17.16) had the highest risk for all-cause mortality^18^. In our study, we demonstrated that the predictive performance of GOLD-R DNAmAge was on par with GrimAge, while surpassing other BAs acceleration in predicting mortality risk in both general populations and cancer survivors. Importantly, this robust performance was achieved with a more streamlined algorithm based on only 34 CpG sites, far fewer than the 1,030 used in GrimAge^6^. In addition, a small set of DNAm CpGs of previous clocks had demonstrated the validity to capture the aging and mortality^15,20^. This parsimony of our GOLD-R DNAmAge highlighted its efficiency and scalability for large-scale or low-coverage applications.

Proteomic aging clocks have traditionally focused on predicting either chronological age^12,21^ or mortality risk^22,23^. For instance, Goeminne et al used Cox models on proteomics data to model survival probability to estimate ProtAge ^22^. A key limitation of chronological age predictors is that their predictive accuracy does not necessarily reflect a capacity to capture biological aging associated with disease and death. Conversely, training mortality-based models requires high-quality longitudinal data with survival profiles, which are heterogeneous across cohorts. Moreover, this field has shifted from organism-level clocks to heterogeneous, organ-specific models for ageotype analysis^10,13,24,25^. For instance, Oh et al. identified brain and immune aging as key predictors of healthspan^13^. However, such models often depended on specific proteins, which may undervalue the role of less-studied, organ-specific proteins. In study, we utilized organ-specific GOLD-R ProtAge models with proteomic specificity^26^. Beyond the brain, we demonstrated that the kidney, stomach, and colon were significant predictors of healthspan. These organs exhibit distinct temporal aging signatures and interact within a multiorgan network^24,27^. Consequently, GOLD-R ProtAge serves as both a high-performance tool for cross-sectional data and a robust metric that quantifies an individual’s proteomic deviation from their expected mortality risk.

Our approach effectively decomposes BA (e.g., DNAmAge) estimation into two components: predicting mortality risk through omics data, and estimating the deviation between predicted mortality risk, and risks estimated from CA. Functionally, it operates like a boosting framework, where CA provides a strong initial estimate, and biomarkers iteratively refine it by capturing residual aging information. This design is highly effective because it leverages known CA variables rather than forcing redundant reconstruction of biomarkers. Furthermore, we defined aging acceleration as the simple difference between BA and CA. In contrast, other methods that directly subtract CA from BA result in a difference score that is inherently negatively correlated with CA, complicating interpretation^14^. Therefore, they defined aging acceleration using the residuals from the regression of BA on CA, a measure that is uncorrelated with CA. In clinical settings, reliably determining effect sizes of CA regression for aging acceleration across diverse populations remains relatively complex.

This study has several limitations, primarily the availability of high-quality DNA methylation data for validation. Future work should include a broader range of health conditions (e.g., geriatric syndromes) to enhance clinical translation and explore integrative multi-omics approaches to further improve mortality risk prediction.

Beyond merely predicting CA, the fundamental aim of aging clocks is to forecast health outcomes^28^. BA predictors have been proposed using longitudinal data derived from epigenetics, clinical-biochemistry, and proteomics^29^. However, owing to differences in study population, objectives, methodologies, and tissue types used across these algorithms, their capacity to capture the true BA status of aging varies considerably. Our findings showed the validity of the GOLD-R algorithm to capture mortality risks with cross-sectional data, which outperformed the age prediction approaches. These results indicate that the GOLD-R framework provides a robust and practical methodology for BA estimation.

## Methods and Materials

### Study Populations

This study utilized data from four sources: EWAS Data Hub, the UK Biobank, the US National Health and Nutrition Examination Survey (NHANES, 1999-2002), and the Health and Retirement Study (HRS, 2016). Statistical data are presented in Table S1-2. EWAS Data Hub is an online resource for collecting and normalizing DNA methylation array data^30^. The UK Biobank is large-scale perspective cohort that collected data from over 500,000 participants across 22 centers in England, Scotland, and Wales. The US NHANES is a nationally representative cross-sectional survey of civilians living in the US. The Health and Retirement Study (HRS) is a longitudinal, nationally representative study that follows approximately 20,000 Americans over age 50, with oversamples of African-American and Hispanic populations. Ethical approval was obtained from the respective review boards for each study (NCHS Ethics Review Board [ERB] for NHANES; North West Multicenter Research Ethics Committee for UK Biobank, ERB of National Institute on Aging and University of Michigan for HRS), and all participants provided written informed consent. This study adheres to the STROBE reporting guidelines for cohort studies.

### Construction of GOLD-R BioAge

The GOLD-R BioAge was constructed using a multi-step, residual-informed framework based on the Gompertz law of mortality. First, the underlying mortality risk (h) was modeled according to the Gompertz law as a function of chronological age (CA):

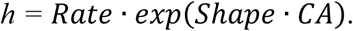

Then, a LASSO regression model with 5-fold cross-validation was then trained to predict the log-transformed mortality risk (𝑙𝑜𝑔 ℎ^) using omics biomarkers (e.g, proteomics, DNAm).

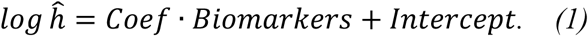

The discrepancy between the methylome-predicted and demographically expected mortality risk was captured as a residual term:

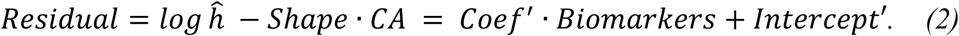

A second LASSO model was developed to predict this residual directly from omics data. The GOLD-R BioAge was then derived by incorporating the predicted residual (PR) back into the Gompertz framework:

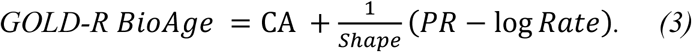

To correct for the underestimation of biological age residuals resulting from the reduced variance of the predicted residuals compared to the original residual risks (RR), we applied a scaling transformation. The predicted residuals were standardized as follows:

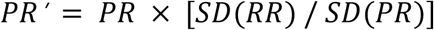

This scaled value was then centered to the mean of the original residuals:

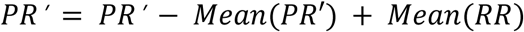

The final GOLD-R BioAge was subsequently calculated using the transformed residual (PR′):

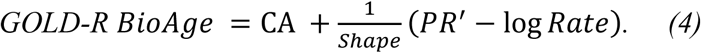

For the NHANES and HRS cohort, where raw DNAm data was unavailable, the residual was derived indirectly from established epigenetic clocks. A composite DNAmAge was calculated from the first five principal components of seven epigenetic clocks regressed on CA, and the residual was computed using formula (2).

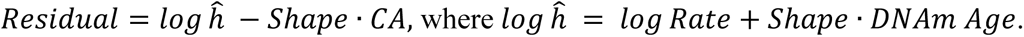

This residual was then predicted using a panel of clinical biomarkers via a LASSO model. The final biological age, termed GOLD-R DNAmCliAge, was derived as the above formula (3, 4). The clinical biomarkers selected for the GOLD-R DNAmCliAge model in NHANES and HRS, along with their coefficients, are provided in **Table S6**.

### Feature Selection of DNAm Data and Model Optimization

DNAm data was obtained from the EWAS Data Hub. The model training analysis selected 7313 adult samples (≥18 years old) to ensure biological relevance to age-related mortality, with Illumina 450K DNAm data (485515 CpGs). For quality control, CpG sites with more than 10% missing rate were removed for analysis and 358149 CpGs were retained. Feature selection was performed by correlation analysis between CpG beta values and chronological age using Pearson correlation. The first Lasso model selected age-correlated CpG sites with absolute Pearson correlation coefficient greater than 0.3. And in second Lasso model, CpG sites were pre-filtered based on their absolute correlation with age, with a sensitivity analysis of correlation thresholds (0.08 to 0.18, **Figure S2**). Feature selection was performed using LASSO regression with 5-fold cross-validation to identify the most predictive CpG subsets.

### Proteomics biomarkers and GOLD-R ProtAge

In UKB (2006-2010), the raw plasma proteomics data (Olink platform) contained 2923 proteins from 53,014 participants. Organ-specific proteins were defined from a previously published resource^26^ Consequently, 1131 proteins from the UKB Olink panel were identified as covering 16 organs. The GOLD-R Proteomic Age (GOLD-R ProtAge) was derived by applying the above residual-informed Gompertz framework, with plasma protein levels as input features. All models were developed and validated using a 5-repeated 5-fold cross-validation scheme, with performance evaluated on mortality and incident disease prediction in the held-out test folds. To ensure optimal model performance, we systematically tuned the correlation thresholds for feature selection in the two-stage LASSO regression (**Figure S4**). Sensitivity analyses assessed the impact of correlation thresholds for feature selection, and model robustness was tested by adding simulated random noise to the test data to mimic cohort stratification.

### Assessment of age-related outcomes

Within the EWAS Data Hub, cancer samples were sourced from The Cancer Genome Atlas (TCGA). These samples included survival information in the form of time-to-event mortality data. In NHANES, mortality data were obtained from the National Death Index (NDI) through December 31, 2019, provided by the Centers for Disease Control and Prevention. Data of mortality status and follow-up duration were available for nearly all participants. In HRS, mortality status and date of death (month and year) were obtained from the publicly available HRS tracker file, which integrates interview records, proxy reports, and linkages to the NDI through July, 2023^31,32^. In UKB, death records were sourced from death certificates held by the National Health Service (NHS) Information Centre (England and Wales) and the NHS Central Register (Scotland) to November 30, 2022. We calculated participants’ time to death from baseline to the date of death, date of loss to follow-up, or last follow-up record. We collected the causes of death, using the International Statistical Classification of Diseases, 10th edition (ICD-10). Cause-specific mortality included mortality of malignant neoplasms, heart disease, cerebrovascular disease, respiratory disease, Alzheimer’s disease, diabetes, and others. In addition, information of incident chronic disease in the UKB, including cancer, myocardial infarction, hypertension, diabetes, stroke, chronic obstructive pulmonary disease (COPD), and dementia, was collected. The health span was defined as the occurrence of seven age-related chronic diseases.

### Benchmark analysis

For comparative analysis with established epigenetic clocks, we utilized the R methylclock package^3^. The DNAmAge function was applied to calculate five prominent epigenetic age measures: Horvath^4^, Hannum^5^, Levine^6^, BNN (Bayesian neural networks)^7^, and Elastic Net^8^ clocks. GrimAge^6^ and DunedinPACE^33^ were calculated through R meffonym and the DunedinPACE package. In NHANES and HRS, epigenetic clocks were downloaded from the official website. Other established phenotypic aging clocks, including Levine’s phenotypic age, KDM age were calculated based on clinical biomarkers, using the R BioAge package. The residual of GOLD-R BioAge was calculated through trained model as a linear combination of biomarkers. The residuals of other clocks were generated through linear regression on CA.

### Statistical Analysis

Cox proportional hazards models were employed to evaluate the association between BA measures and overall survival, adjusting for CA, race, sex, and other covariates (etc., body mass index, smoke, drinking status in UKB). In the UK Biobank, the hazard ratios for organ-specific GOLD-R ProtAge were mutually adjusted using the first three principal components derived from other organ clocks. Survival curves were generated using Kaplan-Meier analysis with cumulative incidence or survival probability as the outcome. Participants were stratified into quartiles based on residuals of epigenetic and phenotypic clocks. All analyses were conducted in R (version 4.2) using the following key packages: biglasso^9^ for LASSO regression, survival for Cox models, survminer for survival visualization, and ggplot2 for graphical representations. Correlation coefficients and p-values were calculated using Pearson correlation with statistical significance defined as p < 0.05.

## Data Availability

Human DNA methylation data was downloaded from EWAS Data Hub (https://ngdc.cncb.ac.cn/ewas/datahub). The data from the NHANES are publicly available at https://www.cdc.gov/nchs/nhanes/index.htm. The data from the HRS are available upon application at https://hrs.isr.umich.edu/. The data from the UK Biobank are available upon application at www.ukbiobank.ac.uk/register-apply. This research was conducted using UK Biobank Resource under Application Number 103791.

## Data and code availability

Human DNA methylation data was downloaded from EWAS Data Hub (https://ngdc.cncb.ac.cn/ewas/datahub). The data from the NHANES are publicly available at https://www.cdc.gov/nchs/nhanes/index.htm. The data from the HRS are available upon application at https://hrs.isr.umich.edu/. The data from the UK Biobank are available upon application at www.ukbiobank.ac.uk/register-apply. This research was conducted using UK Biobank Resource under Application Number 103791. To facilitates application of this approach, we have made the complete workflow publicly available and uploaded the source code to an open-access repository (https://github.com/Jerryhaom/GOLDR-BioAge).

