## Supplementary figures for "A Residual Approach to Estimate Biological Age from Gompertz Modeling"

**Figure S1.** Overview of study datasets and analytical framework.

**Figure S2.** Residual distribution, feature selection and performance of GOLD-R model.

**Figure S3.** Mortality prediction and hazard ratios of epigenetic clocks.

**Figure S4.** Parameter optimisation and performance comparison of GOLD-R ProtAge and its residual model.

**Figure S5.** Correlation analysis of GOLD-R PortAge with chronological age at organismal and organ-specific levels.

**Figure S6.** Analysis of multi-organ GOLD-R ProtAge with disease risk and predictive performance.

**Figure S7.** DNA methylation age (DNAmAge) and residual distributions of GOLD DNAmCliAge.

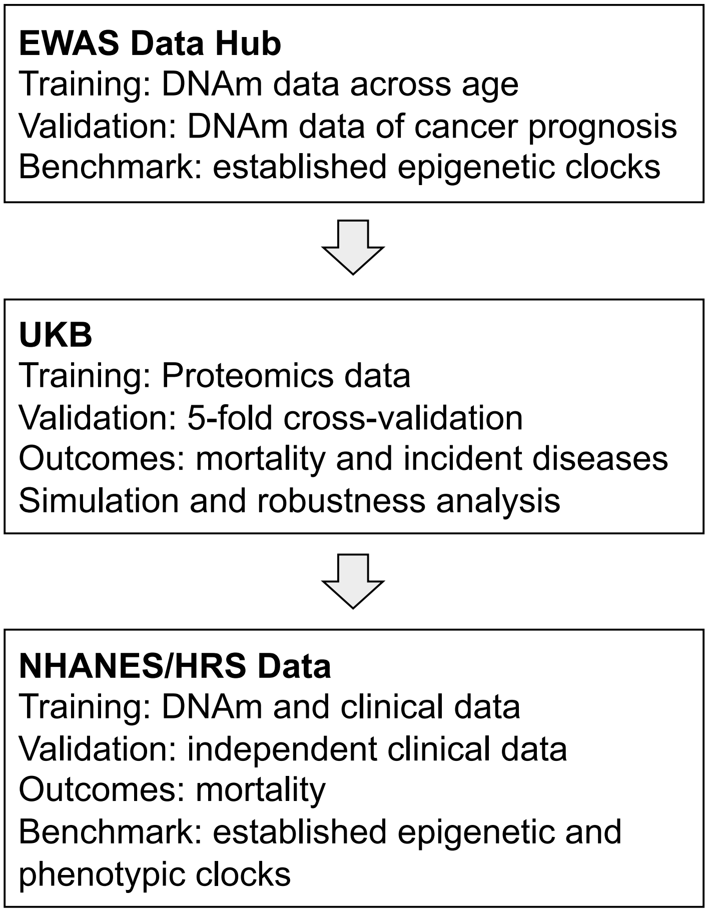


**Figure S1**. Overview of study datasets and analytical framework. UKB: UK Biobank; NHANES: National Health and Nutrition Examination Survey ; HRS: Health and Retirement Study.

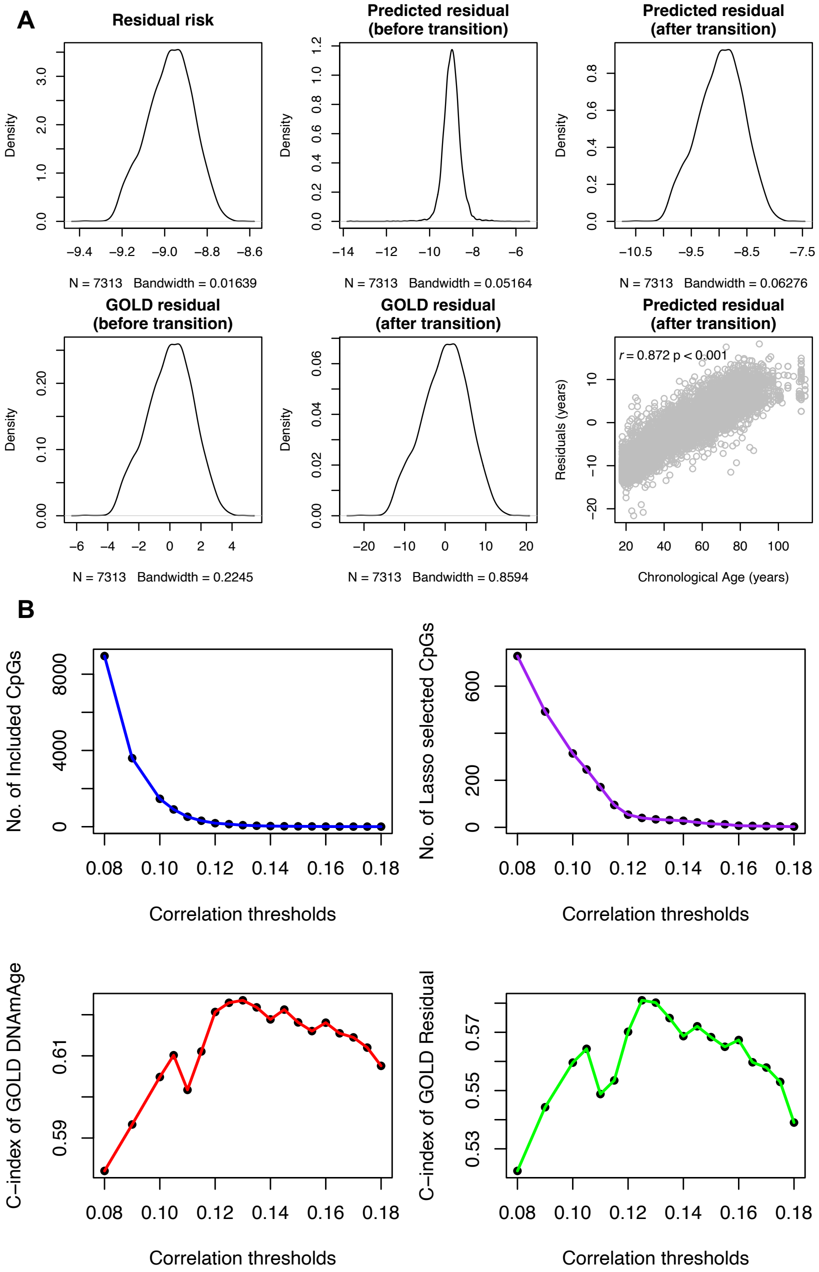


**Figure S2.** Residual distribution, feature selection and performance of GOLD-R model. Figure (A) illustrated distributions of residual risks and unit in years before and after residual transition. The scatter plots showed the correlation of residual with chronological age. (B) demonstrated the relationships of correlation coefficient thresholds with selected CpG sites in LASSO regression (input and enrolled). And the relationships of the thresholds with C-index in mortality prediction in validation dataset.

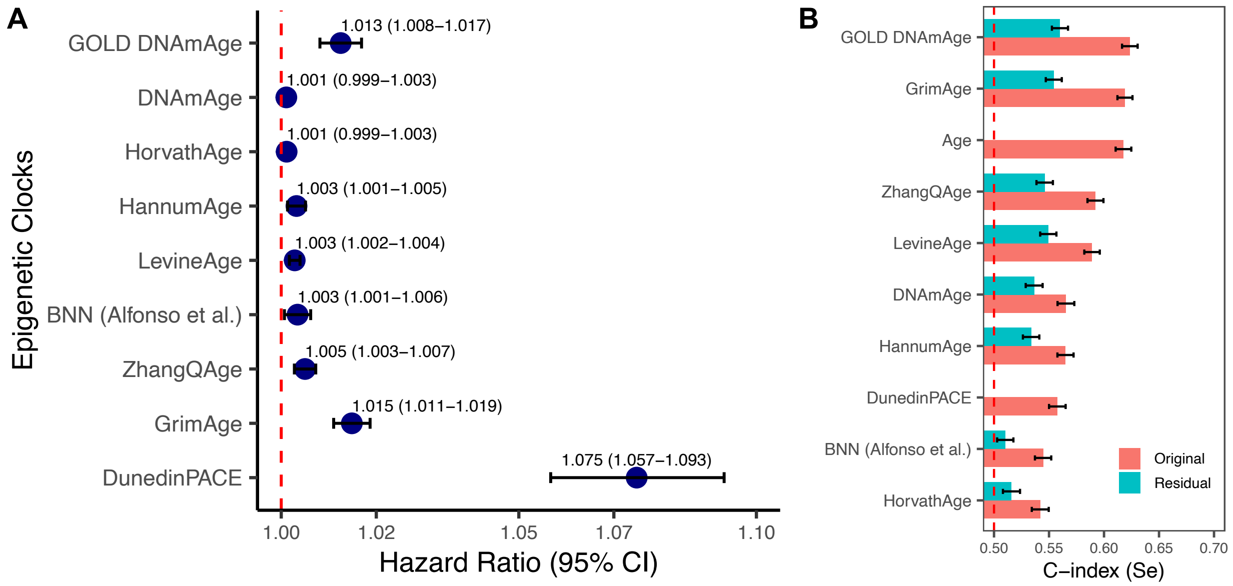


**Figure S3.** Mortality prediction and hazard ratios of epigenetic clocks. The residuals of all clocks were adjusted by the chronological age, through linear regression models. Hazard ratios were calculated after adjusting age, sex, and race in the cox proportional hazard models.

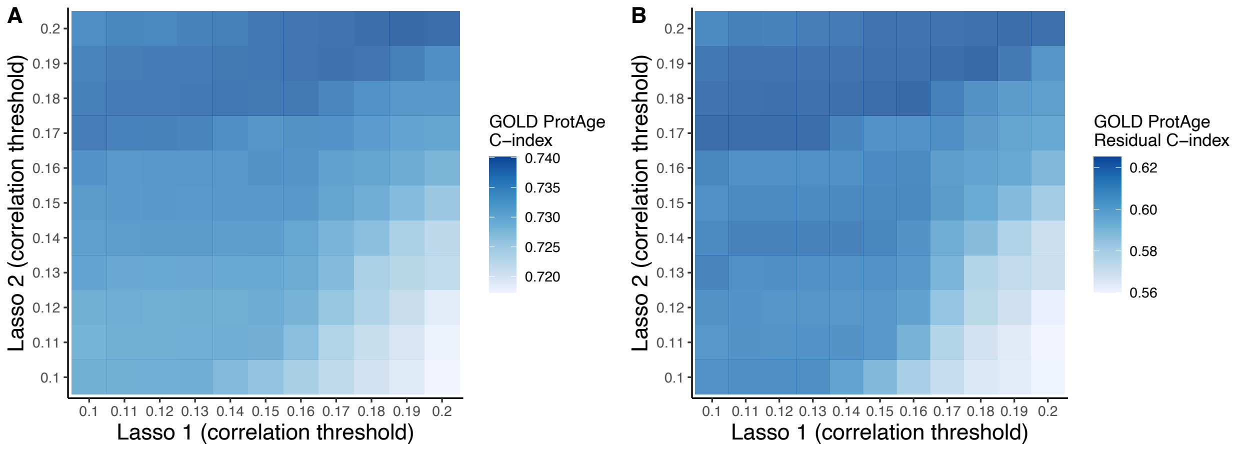


**Figure S4.** Parameter optimisation of GOLD-R ProtAge and its residual. Heatmaps (A) and (B) displayed performance tuning for the original GOLD-R ProtAge model and its residual model respectively. The figure illustrated the predictive performance (C-index) of all-cause mortality regarding to two LASSO regression models (LASSO 1 and LASSO 2) across varying protein correlation thresholds.

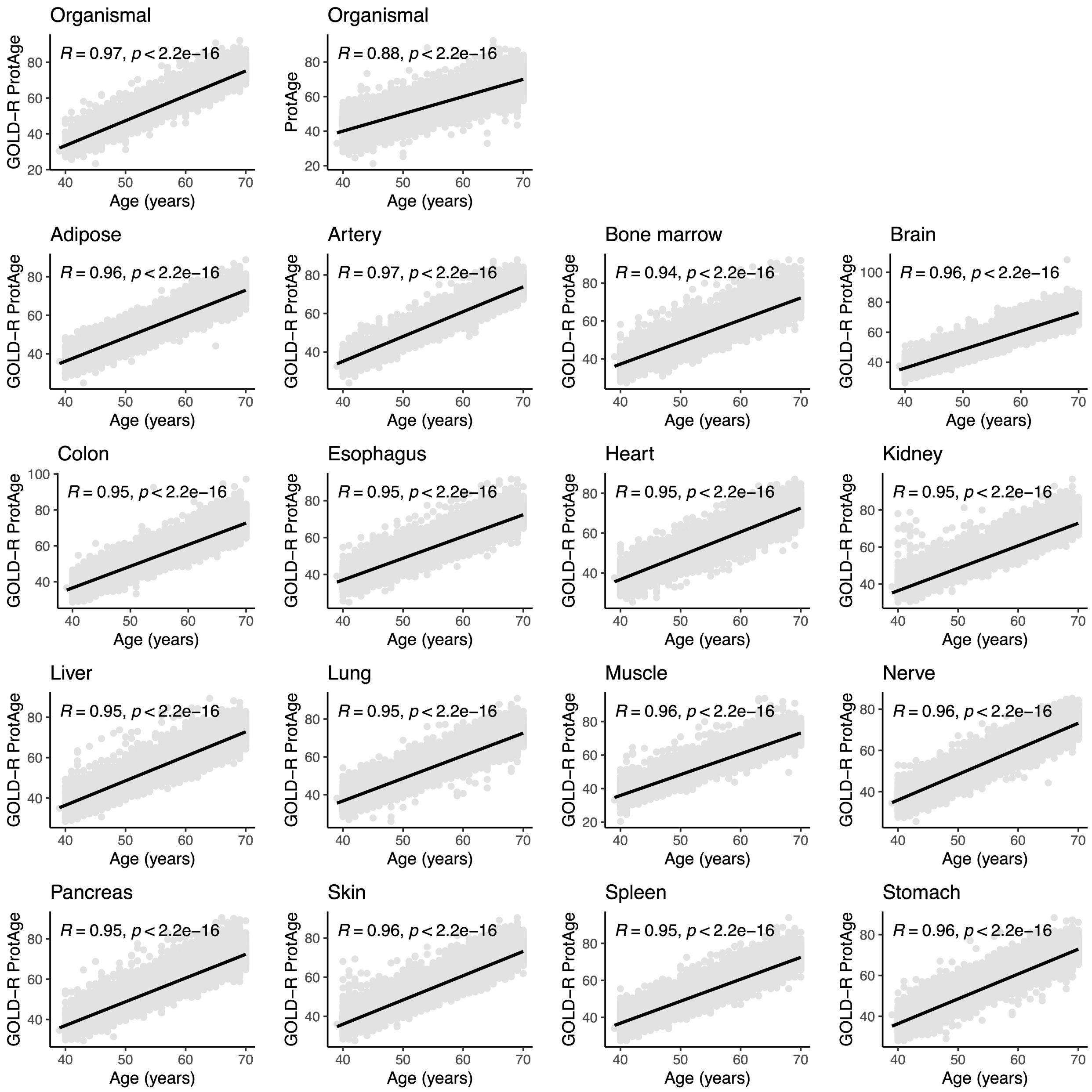


**Figure S5.** Correlation analysis of GOLD-R PortAge with chronological age at organismal and organ-specific levels. Pearson correlation coefficients were shown.

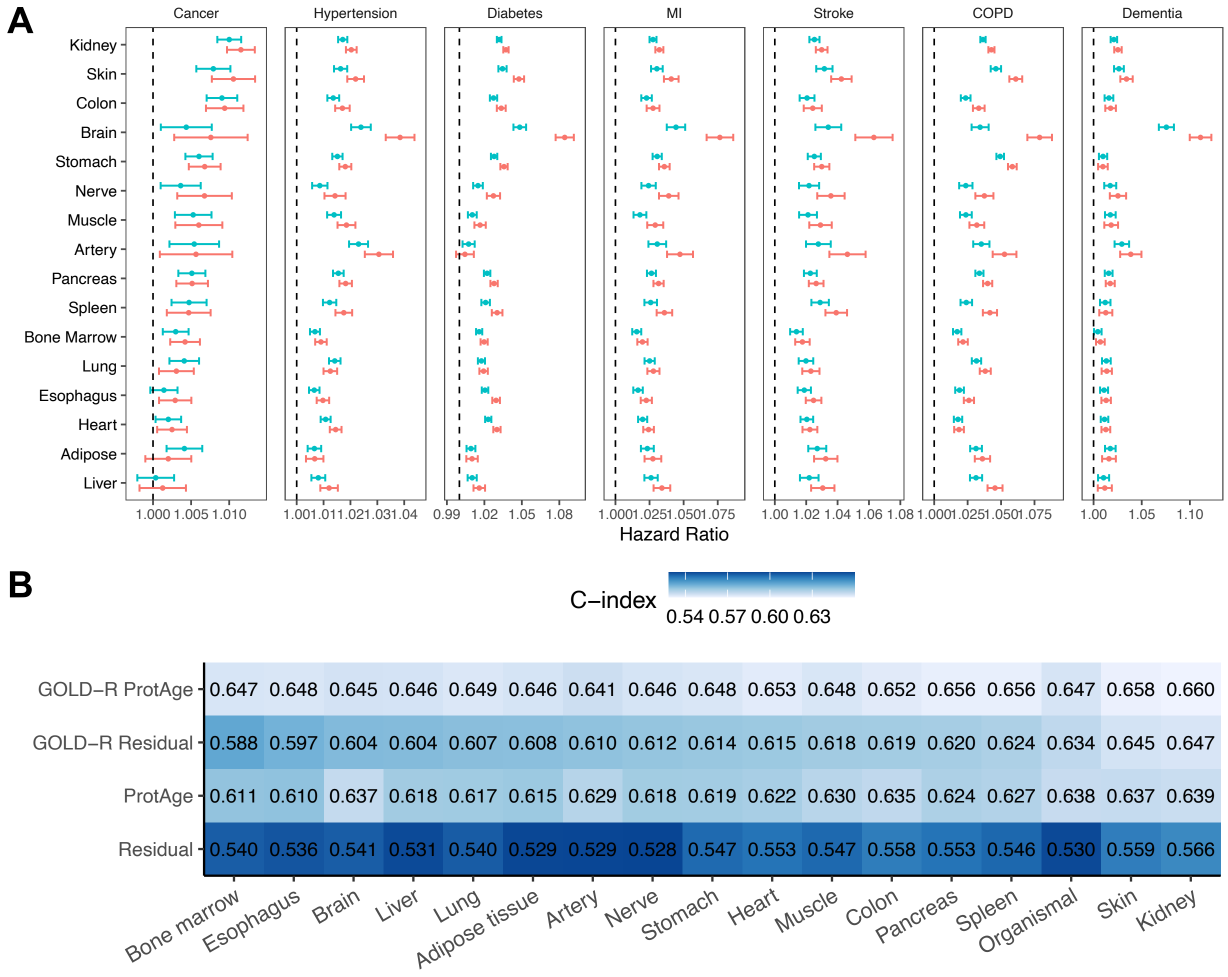


**Figure S6.** Analysis of multi-organ GOLD-R ProtAge with disease risk and predictive performance. Forest plot (A) displayed the adjusted hazard ratios (HR) for seven disease categories (Cancer, Hypertension, Diabetes, MI, Stroke, COPD, Dementia) across different organs. Hazard ratios were adjusted for age, sex, race, BMI, smoking and drinking status. The heatmap (B) presented the clinical predictive performance metrics (C-index) for disease incidence prediction across various organs using different biological age models (GOLD-R ProtAge, GOLD-R Residual, ProtAge, Residual). MI: Myocardial Infarction. COPD: Chronic Obstructive Pulmonary Disease.

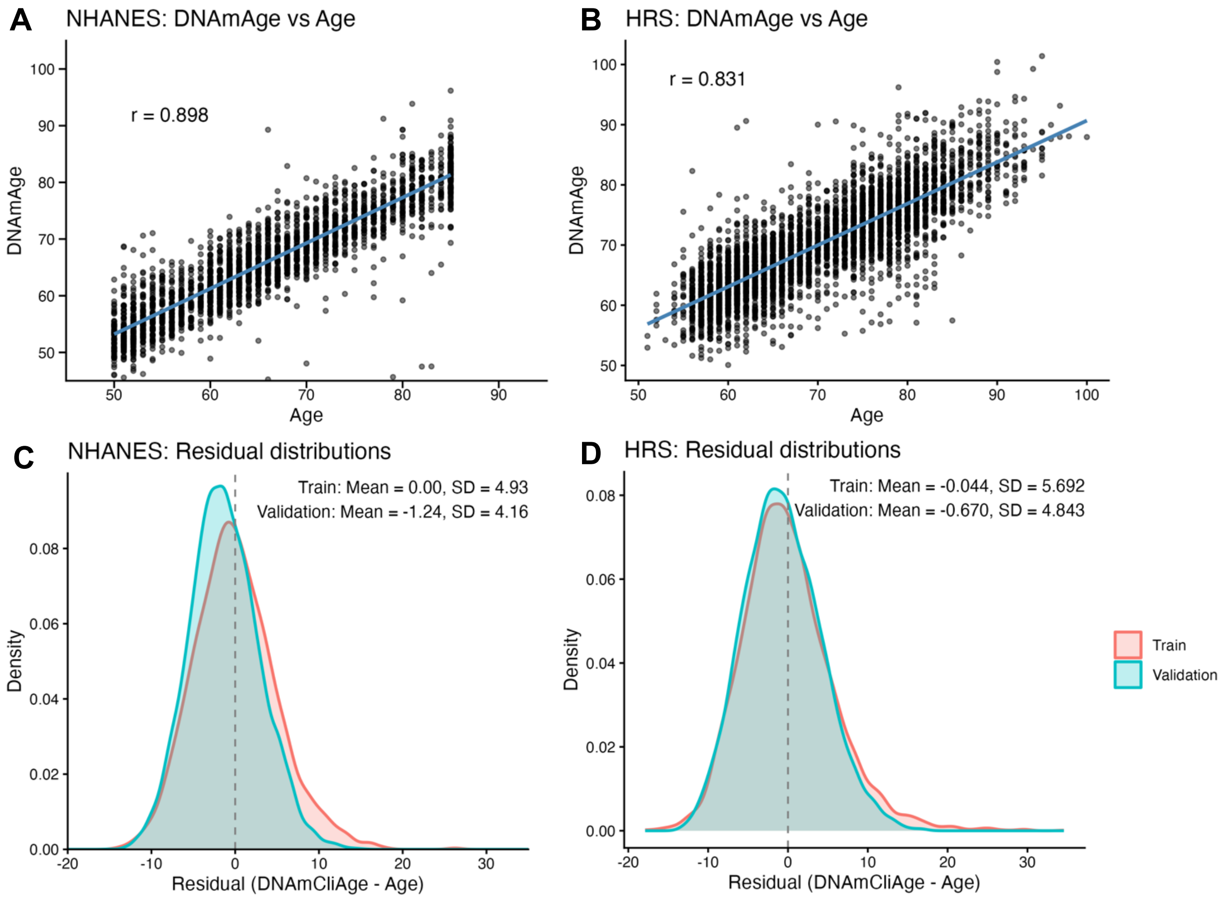


**Figure S7.** DNA methylation age (DNAmAge) and residual distributions of GOLD DNAmCliAge. Scatter plots A and B showed the scatter plots of DNAmAge and age constructed from the first 5 PCs and the Pearson correlation coefficient (r) for the NHANES and HRS datasets, respectively. Panels C and D showed the distribution density of residuals (the difference between predicted age and actual age) in the two cohorts. SD: Standard deviation.
